# Door-to-Valve Intervention Time and Clinical Outcomes in Acute Decompensated Severe Aortic Stenosis: Systematic Review and Meta-Analysis

**DOI:** 10.64898/2026.11.03.26367226

**Authors:** Jeff R Ma, Mark Gerges, Anoop N Koshy, William Wilson, Andrew J Taylor, William Chan

## Abstract

**Background:** Acute decompensated aortic stenosis (ADAS) carries high mortality, but the optimal timing of valve intervention is unknown.

**Objective:** To assess the association between time to valve intervention (transcatheter aortic valve replacement [TAVR] or balloon aortic valvuloplasty [BAV]) and clinical outcomes in ADAS.

**Methods:** Embase, Ovid MEDLINE and CENTRAL were searched to May 15, 2026. Hazard ratios (HRs) for delayed versus earlier intervention were pooled using random-effects models with modified Hartung-Knapp inference. Unadjusted short-term mortality risk ratios (RRs) were pooled using the Mantel-Haenszel method.

**Results:** Eight observational cohort studies (n=6,044) met inclusion criteria: six TAVR (n= 5,986) and two BAV (n=58) studies. Patients were approximately 79 years old, and 63% were male. Early-intervention thresholds ranged from <24-48 hours (BAV) to ≤22 days (TAVR) from admission. Delayed intervention was not significantly associated with long-term all-cause mortality (HR 1.52, 95% CI 0.46-5.00; p=0.269; I^2^=58.2%) or mortality per five-day delay (HR 1.09, 95% CI 0.34-3.43; p=0.530; I^2^=86.7%). Crude short-term mortality was higher with delayed intervention (RR 4.96, 95% CI 1.17-20.92; p=0.029; I^2^=0.0%), although this association was not robust to sensitivity analyses. Most studies were at serious risk of bias, with substantial heterogeneity in clinical severity, intervention timing, and treatment modality.

**Conclusion:** Point estimates favour earlier intervention, but current observational data do not establish a mortality benefit for an optimal door-to-valve interval, and existing study designs are biased against early treatment. Prospective and randomized-controlled trials on optimal valve intervention timing following ADAS are needed.

## BACKGROUND

Severe aortic stenosis (AS) affects approximately 4% of adults aged ≥75 years, and is projected to double by 2050 [1]. Access to timely valve replacement remains challenging, with almost one in four patients failing to receive planned intervention within three months of meeting guideline criteria [2, 3]. Prolonged waiting times have been associated with clinical deterioration, acute decompensation and mortality [4].

Acute decompensated aortic stenosis (ADAS) represents a particularly high-risk clinical presentation, characterised by acute heart failure, haemodynamic instability, or cardiogenic shock requiring hospitalisation and consideration of urgent valve intervention. These patients experience substantial morbidity, prolonged hospitalisation and mortality exceeding 20% [5]. Although transcatheter aortic valve replacement (TAVR) is increasingly used and technically feasible in this setting [6], optimal intervention timing remains uncertain. Earlier relief of valvular obstruction may prevent progressive myocardial and end-organ dysfunction, but must be balanced against clinical stabilisation and pre-procedural assessment.

These challenges have prompted increasing interest in door-to-valve time, defined as the interval from hospital presentation to relief of valvular obstruction through TAVR or balloon aortic valvuloplasty (BAV), as a potentially modifiable systems-of-care metric [7]. Although contemporary interventional guidelines recommend early intervention within 90 days for severe AS [8], they provide no specific time-to-intervention targets for patients presenting with acute decompensation. Furthermore, heterogeneous definitions of acute decompensation, intervention timing and clinical outcomes across existing studies limit the development of evidence-based treatment pathways.

We therefore conducted a systematic review and meta-analysis to evaluate the association between timing of valve intervention and clinical outcomes in patients with ADAS, and to assess the current evidence supporting door-to-valve time as a potential quality-of-care metric.

## METHODS

Embase, Ovid MEDLINE and the Cochrane Central Register of Controlled Trials were searched from inception to 15 May 2026 for full-text randomised and observational studies of adults with ADAS comparing early versus delayed TAVR or BAV. The search combined Medical Subject Headings and free-text terms including “acute decompensated aortic stenosis”, “acute valve syndrome”, “transcatheter aortic valve replacement”, “balloon aortic valvuloplasty” and “timing” with syntax adapted to each database. Early and delayed status was defined as per individual study. Two independent reviewers screened abstracts and full texts (JRM, MG), supplemented by backward and forward citation searching. Disagreements were resolved by a third independent reviewer (WC). For studies meeting eligibility criteria, data were extracted by JRM and MG using a standardised form. Where populations overlapped, the most complete dataset was retained for quantitative analysis. Overall and domain-level risk of bias were assessed using ROBINS-I Version 2. The study followed PRISMA 2020 guidelines. The protocol was prospectively registered with PROSPERO (CRD420261376181). Ethics approval was exempt because all data were derived from previously published studies.

The primary outcome was all-cause mortality according to study-defined early versus delayed intervention. Secondary exploratory outcomes were all-cause mortality per five additional days to valve intervention and crude short-term mortality - prioritising 30-day over in- hospital mortality where both were available.

Continuous variables were summarised as mean ± standard deviation or median with interquartile range (IQR), as appropriate, and categorical variables as counts and percentages, drawn from available data. Hazard ratios (HRs) were pooled on the log scale by generic inverse-variance weighting using restricted maximum-likelihood (REML) random-effects models with modified Hartung-Knapp confidence intervals. Where HRs were unavailable in the manuscript, approximate individual participant data were reconstructed from digitised Kaplan-Meier curves using the Guyot algorithm, incorporating reported risk sets and event totals. HRs were harmonised for direction. Continuous estimates were standardised to five-day increments by rescaling log HRs and standard errors (SEs), assuming a log-linear association. Missing SEs were derived from confidence intervals or reported two-sided p-values, assuming a Wald test of the same coefficient. Unadjusted mortality risk ratios (RR) were pooled directly from 2×2 tables using a common-effect Mantel-Haenszel method, without continuity correction of the pooled counts. Where zero events were identified in one cell, 0.5-corrected REML random-effects estimates with modified Hartung-Knapp inference were examined. Exploratory safety analyses used common-effect Mantel-Haenszel risk ratios without continuity correction. Heterogeneity was summarised using τ² and I². Given the small study numbers, publication-bias tests and meta-regression were not undertaken. Analyses were performed in R 4.5.1 with metafor 4.8-0.

## RESULTS

Of 431 studies identified from searched databases and citation chasing, 51 duplicates were removed and 380 underwent title and abstract screening. After exclusion of 369 studies, eleven full texts were assessed for eligibility and eight studies were included in the qualitative synthesis (Figure 1). All eight contributed to at least one quantitative synthesis; seven contributed to the three principal mortality analyses.

**Figure 1.**
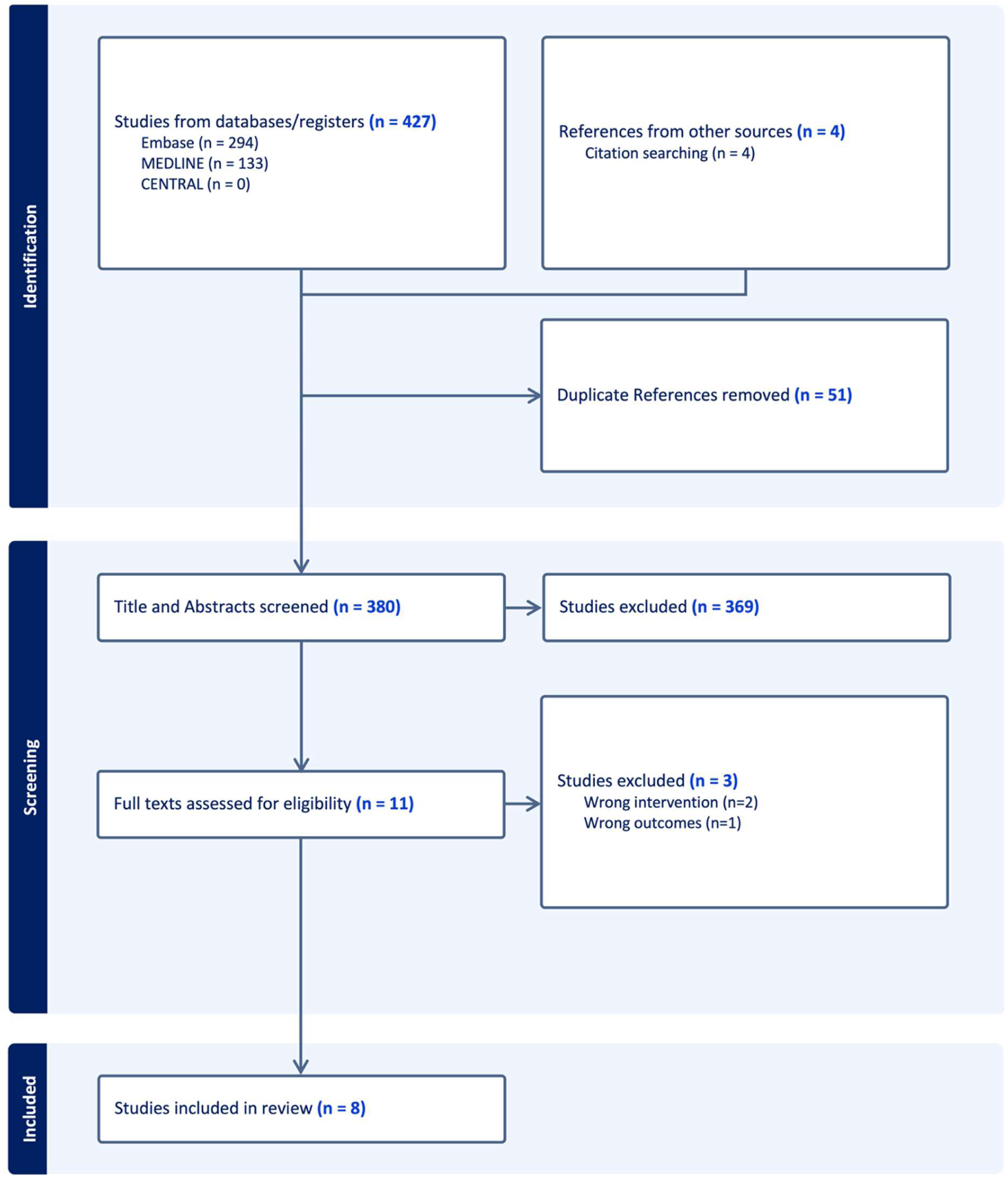
PRISMA diagram. CENTRAL, Cochrane Central Register of Controlled Trials.

Eight observational cohorts comprising 6,044 patients were included, with six TAVR cohorts (n=5,986) and two BAV cohorts (n=58). Median study-level age was 78.7 (75.2-82.7) and 63.4% (3,657/5,767) were male. Hypertension was present in 85.8% (4,856/5,657), and 42.8% (2,588/6,043) had diabetes (Table 1). The median study-level echocardiography estimates were 0.62 cm^2^ (range, 0.50-0.70) for aortic valve area, 42 mmHg (range, 40-46) for mean transvalvular gradient, and 42.9% (range, 32.4-52.8) for left ventricular ejection fraction (LVEF).

**Table 1A.** Study design, populations and timing definitions. HR harmonised such that >1 indicates greater hazard with delay. Where multiple HRs are reported, the adjusted HR with the longest follow-up was selected. ADAS, acute decompensated aortic stenosis; AHF, acute heart failure; AKI, acute kidney injury; AS, aortic stenosis; BAV, balloon aortic valvuloplasty; CP cardiopulmonary; CS, cardiogenic shock; CXR, chest radiograph; HF, heart failure; HR, hazard ratio; IV, intravenous; MCS, mechanical circulatory support; TAVR, transcatheter aortic valve replacement; TVT, Transcatheter Valve Therapy

| Study | Setting and design | Relevant cohort | Procedure | ADAS population | Timing comparison | Relevant outcome | Hazard ratio |
|---|---|---|---|---|---|---|---|
| McKenna 2025 **[22]** | UK; single centre; retrospective; 2015–2022 | N=276; early 150, delayed 126 | TAVR | Dyspnoea 208 (75%), syncope 68 (25%); 8 (2.9%) requiring CP support | 1. ≤22 vs >22 days from admission.<br>2. Continuous admission-to-TAVR delay per 5 days | HF hospitalisation or all-cause mortality | 1. Unadjusted HR 1.48 (95% CI 1.09-2.03; p=0.013)<br>2. Adjusted HR 1.09 (95% CI 1.04-1.16; p=0.001) |
| Lux 2021 **[23]** | The Netherlands; single centre; retrospective; 2013–2018 | N=53 | TAVR | Dyspnoea 73.6%, angina 30.5%, syncope 22.6% | 1. Continuous admission-to-TAVR delay per 1 day | All-cause mortality | Unadjusted HR 1.037, p=0.003 |
| Patel 2023 **[17]** | UK; single centre; prospective non-randomised cluster study; 2019–2022 | N=83; early 25, delayed 58 | TAVR | Syncope, acute pulmonary oedema, dyspnoea at rest; cardiogenic shock excluded | 1. 10 (6-12) vs 22 (15-30) days from admission | Composite of all-cause mortality or AKI (Stage >2+) | Unadjusted HR 2.35 (95% CI 0.62–8.84; p=0.208) |
| Goel 2023 [6] | USA; national TVT registry; retrospective; 2015–2022 | N=5,006 | TAVR | 1. CS coding;<br>2. Pre-procedural inotropes or MCS; and/or<br>3. Cardiac arrest within 24 hours pre-TAVR | 1. ≤5 vs >5 days from admission | All-cause mortality | Adjusted HR 1.10 (95% CI 0.97-1.23; p=0.149) |
| Debry 2018 **[19]** | France; two centres; retrospective; 2011–2016 | N=31; early 18, delayed 13 | BAV | CS defined as cardiac index <2.2L/min/m<sup>2</sup> + pulmonary congestion resistant to high dose IV loop diuretic and peripheral hypoperfusion, (e.g. oliguria <30ml/hr, lactate >2.0mmol/L) together with hypotension | 1. ≤48 vs >48 hours from inotrope initiation in hypotensive shock | All-cause mortality or recurrent CS related to AS | Reconstructed HR 1.97 (95% CI 0.87-4.44) |
| Buchwald 2001 **[20]** | Germany; single centre; retrospective; 1989–1998 | N=14; early 4, delayed 10 | BAV | CS defined as pulmonary oedema on exam, CXR on admission requiring inotropes, mechanical ventilation or both | 1.<24 vs >48 hours from shock onset | All-cause mortality | N/A |
| Kaewkes 2021 **[18]** | USA; single centre; retrospective; 2013–2017 | N=139; early 43, delayed 96 | TAVR | AHF by Framingham criteria and confirmed by cardiologist; hypotension excluded | 1. <60 vs ≥60 hours from emergency department arrival | All-cause mortality | Adjusted HR 2.38 (95% CI 0.99-5.88; p=0.053) |
| Elbaz-Greener 2019 **[24]** | Canada; 10 centres; retrospective; 2010–2016 | N=429 | TAVR | Declining medical status during concurrent hospitalisation | 1. Continuous referral-to-TAVR delay per 1 day | All-cause mortality | Adjusted HR 1.00 (95% CI 0.9-1.01; p=0.40) |

**Table 1B.** Baseline clinical characteristics. Data are mean ± standard deviation, median [interquartile range], or n/N (%). AVA, aortic valve area; LVEF, left ventricular ejection fraction; NR, not reported; PCI, percutaneous coronary intervention. * McKenna was excluded from aggregate demographic percentages because of possible overlap with Patel.

| **Study** | **Age, years** | **Male, n/N (%)** | **Hypertension, n/N (%)** | **Diabetes, n/N (%)** | **Prior PCI, n/N (%)** | **LVEF, %** | **AVA, cm²** |
|---|---|---|---|---|---|---|---|
| McKenna 2025 * | 84 [79–88] | 176/276 (63.7) | 144 (52.5) | 72 (26.2) | Within 90 days:<br>14/276 (5.1) | 55 [37–58] | 0.7 [0.5–0.8] |
| Lux 2021 | 79 [73–85] | 30/53 (56.6) | NR | 17/53 (32.1) | NR | <30% in 30.2% | NR |
| Patel 2023 | Early: 84 ± 7<br>Delayed: 82 ± 7 | 47/83 (56.6) | 60/83 (72.3) | 28/83 (33.7) | NR | Early: 47 [33–55]<br>Delayed: 53 [37–55] | Both: 0.6 [0.5–0.8] |
| Goel 2023 | 75.56 ± 11.03 | 3,229/5,005 (64.5) | 4,279/5,006 (85.5) | 2,185/5,005 (43.7) | 1,598/5,003 (31.9) | 39.79 ± 17.64<br>(n=4,975) | 0.695 ± 0.319<br>(n=4,639) |
| Debry 2018 | 77.3 ± 8.1 | 33/44 (75.0) | NR | 14/44 (31.8) | NR | 30 ± 14 | 0.61 ± 0.17 |
| Buchwald 2001 | 74.1 ± 11.4 | 7/14 (50.0) | NR | 3/14 (21.4) | NR | NR | 0.38 ± 0.09 |
| Kaewkes 2021 | 82.2 ± 9.6 | 74/139 (53.2) | 122/139 (87.8) | 58/139 (41.7) | 25/139 (18.0) | 46.0 ± 17.7 | 0.62 ± 0.20 |
| Elbaz-Greener 2019 | 81.3 ± 8.9 | 237/429 (55.2) | 395/429 (92.1) | 211/429 (49.2) | 121/429 (28.2) | NR | NR |

**Table 1C.** Hospital stay, follow-up and outcomes. Values in square brackets are interquartile ranges. Mortality and follow-up are reported at study-specific time points. IQR, interquartile range; ICU, intensive care unit; NR, not reported; TAVR, transcatheter aortic valve replacement.

| Study | Time to intervention | Hospital stay, days | Mortality, early vs delayed | Follow-up | Risk of Bias |
|---|---|---|---|---|---|
| McKenna 2025 | 22 [13–32] days overall; early 14 [10–19], delayed 33 [26–42] | Early 17 [11–22], delayed 33 [27–46] | 139/276 (50.4%) | Median 4.6 years (IQR 3.6–5.7) | Serious |
| Lux 2021 | 18 [10–28] days overall | NR | 30-day: 11.3%<br>1-year: 28.3% | 30 days and 1 year | Serious |
| Patel 2023 | 15 days overall; early 10 [6–12] days, delayed 22 [15–30] days | Early 13 [8–18], delayed 24 [18–33] | 0/25 (0%) vs 4/58 (6.9%) | 30 days | Moderate |
| Goel 2023 | 5 [2–9] days overall | NR | 30-day: 11.6% vs 14.3%<br>1-year: 26.5% vs 33.3% | 30 days and 1 year | Moderate |
| Debry 2018 | 4.1 ± 2.9 days from ICU admission and 3.2 ± 3.8 from inotrope initiation overall | Overall 18.3 ± 12.4 days | 1-month: overall 21/44 (47.7%)<br>1-year: 54% vs 86%; overall 31/44 (70.5%) | 1 month and 1 year | Serious |
| Buchwald 2001 | Early 8–18 hours; delayed 48 hours–8 days after shock onset | NR | In-hospital: 0/4 (0%) vs 10/10 (100%)<br>1-year: 0/4 (0%) vs 10/10 (100%) | In-hospital and 1 year | Serious |
| Kaewkes 2021 | 5 [1.5–7.0] days overall | Early 4 [3–6], delayed 11 [7–16] | 30-day: 2/43 (4.7%) vs 9/96 (9.4%)<br>2-year: 6/43 (14.0%) vs 33/96 (34.4%) | 30 days and 2 years | Serious |
| Elbaz-Greener 2019 | 36 [14–95] days overall | Overall post-TAVR 15.0 ± 25.2 | 49/429 (11.4%) | 30 days | Serious |

Previous PCI was reported in 31.3% (1,744/5,571) of participants across three cohorts. McKenna et al reported PCI within 90 days before TAVR in 5.1% (14/276), including 4/150 in the early-intervention group and 10/126 in the delayed-intervention group. Goel et al reported concomitant PCI in 1.5% (76/5,006) of TAVR recipients. Valve-in-valve TAVR accounted for 12.2% of procedures across the two cohorts reporting this variable. The median reported study follow-up was 1 year. Overall risk of bias was moderate in two studies and serious in six.

### Primary outcome

Three studies contributed to the primary synthesis of all-cause mortality at 1 or 2 years. Delayed intervention was not significantly associated with increased all-cause mortality (HR 1.52, 95% CI 0.46-5.00; p=0.269; I^2^=58.2%; Figure 2). Leave-one-out sensitivity analyses yielded pooled HRs ranging from 1.32 to 2.22, with all confidence intervals crossing 1. In contrast, a common-effect sensitivity analysis demonstrated a modest but statistically significant association (HR 1.13; 95% CI 1.003-1.27; p=0.045; I^2^=59.6%), highlighting sensitivity to modelling assumptions.

**Figure 2.**
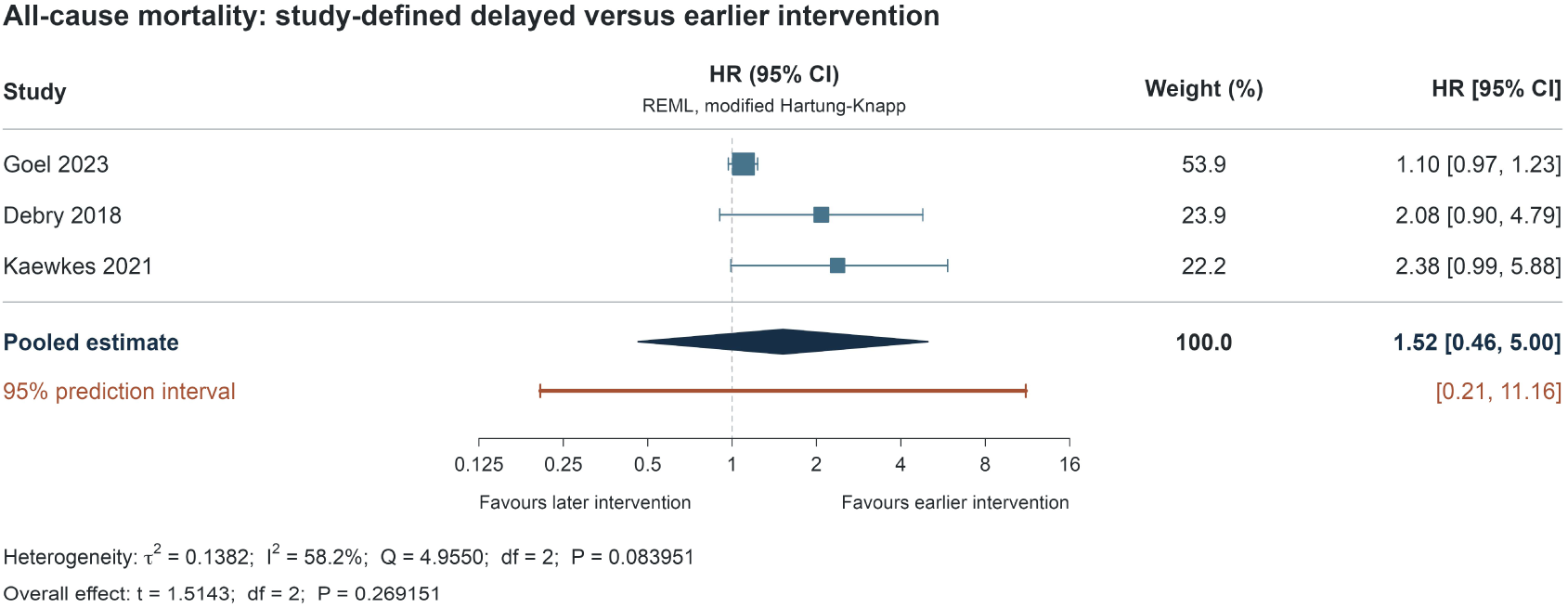
All-cause mortality at 1-2 years according to study-defined timing of delayed versus early intervention. Goel and Kaewkes provide adjusted TAVR HRs. Debry provides unadjusted BAV HR. CI, confidence interval; HR, hazard ratio; REML, restricted maximum-likelihood; TAVR, transcatheter aortic valve replacement.

### Secondary exploratory analysis

Two studies evaluated the association between time to intervention as a continuous exposure and all-cause mortality. No statistically significant association was observed per five-day delay to TAVR (HR 1.09, 95% CI 0.34-3.43; p=0.530; I^2^=86.7%; Figure 3). An expanded analysis incorporating McKenna et al’s composite outcome of death or heart failure admission similarly demonstrated no significant association (HR 1.08 per five additional days, 95% CI 0.87-1.34; p=0.261; I2=83.3%; Figure S1).

**Figure 3.**
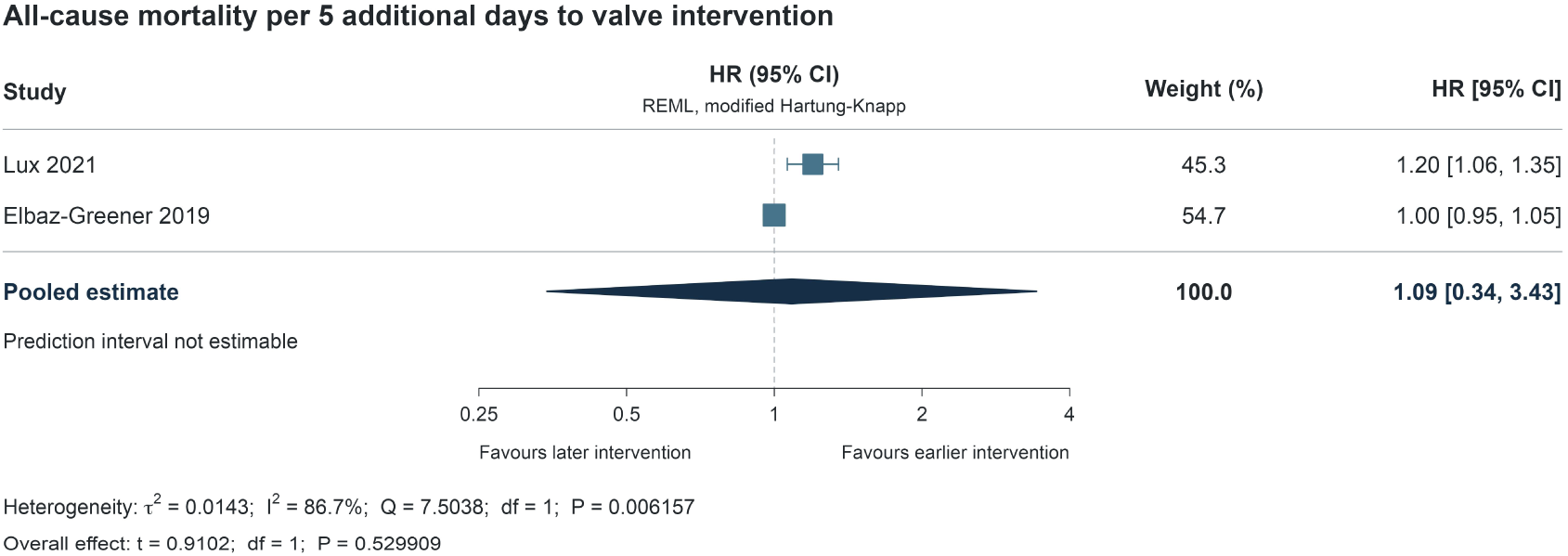
All-cause mortality per five additional days to intervention. CI, confidence interval; HR, hazard ratio; REML, restricted maximum-likelihood.

Three studies contributed crude short-term mortality data. Deaths occurred in 2/72 patients in the early-intervention groups and 23/164 in the delayed-intervention groups. Delayed intervention was associated with significant higher crude short-term mortality (RR 4.96, 95% CI 1.17-20.92; p=0.029; I^2^=0%; Figure 4). However, this association was not statistically significant in a continuity-corrected random-effects sensitivity analysis (RR 3.09, 95% CI, 0.23–41.43; p=0.202; I^2^=0%). Similarly, statistical significance was lost following exclusion of either Buchwald et al. (RR 2.89, 95% CI 0.67–12.45; p=0.155; I^2^=0%) or Patel et al. (RR 4.08, 95% CI 0.96–17.40; p=0.057; I^2^=20.8%).

**Figure 4.**
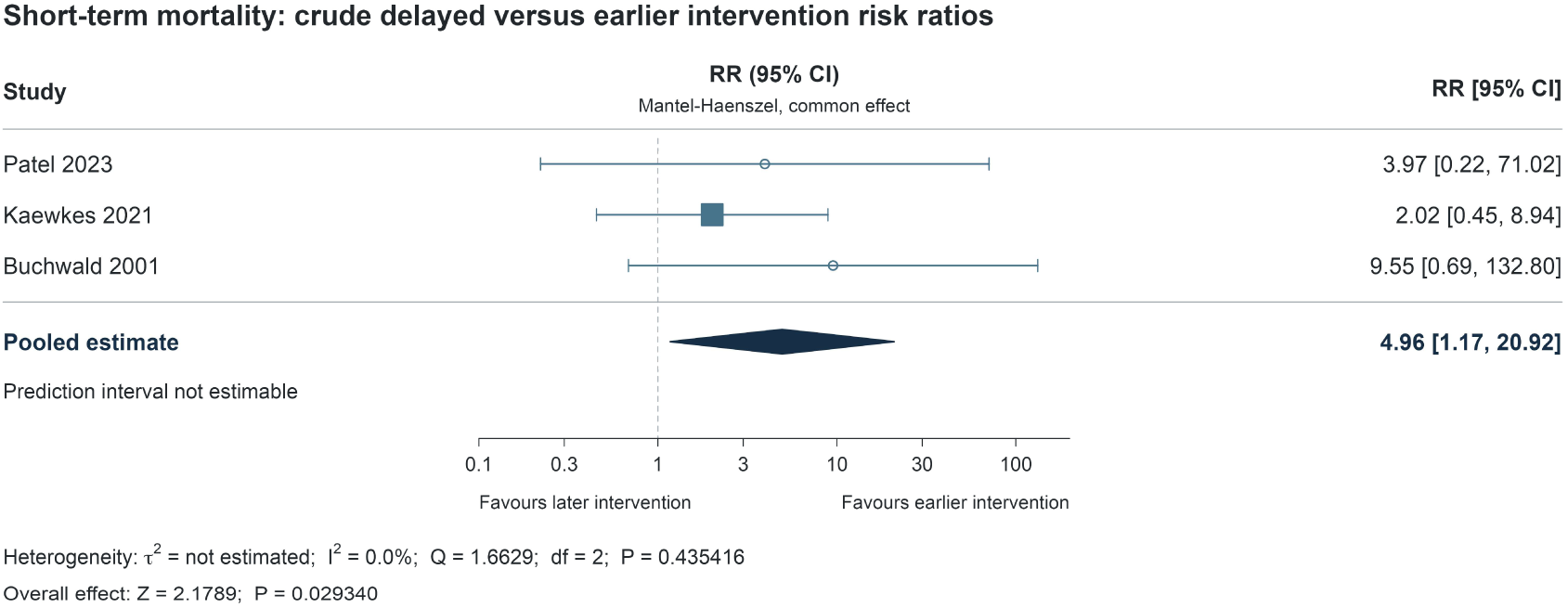
Crude short-term mortality. Common-effect Mantel-Haenszel RR without continuity correction to the pooled estimate. CI, confidence interval; RR, risk ratio.

Supplementary analyses demonstrated no statistically significant association between delayed intervention and study-defined mortality or death-containing composite outcomes (HR 1.40, 95% CI 0.84–2.32; p=0.126; I²=60.2%; Figure S2). Similarly, no significant association was observed for 30-day mortality restricted to TAVR recipients (RR, 2.68; 95% CI, 0.62–11.59; P=0.188; I^2^=0%; Figure S3) or total hospital length of stay (mean difference, delayed minus early intervention, 12.77 days; 95% CI, −61.34 to 86.89; P=0.273; I²=98.0%; Figure S4).

### Procedural safety endpoints

Timing-stratified procedural safety data were inconsistently reported. Among available data, there were no statistically significant differences between the early- and delayed-intervention groups in major bleeding (7 [4.0%] in early versus 4 [2.2%] in delayed; p=0.284), pacemaker implantation (21 [12.0%] versus 13 [7.1%]; p=0.131), major vascular complications (8 [4.6%] versus 5 [2.7%]; p=0.506), minor vascular complications (10 [5.7%] versus 8 [4.3%]; p=0.415), or stroke (3 [1.7%] versus 2 [1.1%]; p=0.798).

## DISCUSSION

This systematic review and meta-analysis identified three main findings. First, delayed valve intervention was not significantly associated with all-cause mortality in the primary analysis, although effect estimates generally favoured earlier intervention. An association with increased crude short-term mortality was observed but was not robust to sensitivity analyses. Second, procedural safety seemed comparable, although estimates were limited by sparse events and incomplete reporting. Third, heterogeneity in the ADAS population and timing definitions precluded identification of an optimal treatment interval. To our knowledge, this is the first systematic review and meta-analysis evaluating door-to-valve time in ADAS. Collectively, these findings highlight an important evidence gap: although expedited intervention is physiologically plausible and potentially modifiable at a systems level, current evidence is insufficient to establish a mortality benefit or define an optimal door-to-valve target.

Chronic pressure overload promotes concentric hypertrophy and diastolic dysfunction, increasing dependence on adequate filling. Superimposed stressors - including volume shifts, ischaemia or tachyarrhythmia such as atrial fibrillation - may overwhelm this limited reserve, elevate left atrial and pulmonary vein pressures, and precipitate congestion despite initially preserved, compensated systemic perfusion [9]. With further deterioration, inadequate cardiac output produces organ hypoperfusion, while subendocardial ischaemia secondary to increased wall stress and myocardial oxygen demand may culminate in cardiogenic shock [10]. These presentations broadly correspond to patients at risk of shock or with emerging haemodynamic instability (Society for Cardiovascular Angiography and Interventions [SCAI] stages A-B), through to established and progressive shock (stages C-E) [11], suggesting that the clinical importance of intervention timing may differ according to the severity and trajectory of decompensation.

Although not statistically significant, the consistent direction of the pooled mortality estimates supports prospective evaluation of door-to-valve time as a quality-of-care target. Door-to-balloon and door-to-needle times demonstrate how treatment intervals can organise services around timely interventions in ST-elevation myocardial infarction (STEMI) and acute ischaemic stroke [12, 13]. Similarly, the emerging “door-to-support” paradigm in STEMI complicated by cardiogenic shock (CS) emphasises early haemodynamic stabilisation and mechanical circulatory support – ideally achieving left ventricular unloading within 90 minutes [14–16]. Although these conditions differ fundamentally from ADAS, they illustrate the potential value of coordinated, time-sensitive care pathways. In ADAS, TAVR or BAV relieves the fixed obstruction driving haemodynamic compromise, providing a disease-specific target for an expedited pathway. However, any proposed door-to-valve target must balance the potential benefits of earlier intervention against the need for clinical stabilisation and appropriate procedural assessment.

Door-to-valve time captures modifiable delays across diagnosis, referral, inter-hospital transfer, pre-procedural imaging, and procedural access. The Rapid Assessment and Treatment in Decompensated Aortic Stenosis (ASTRID-AS) study provided a template pathway which included early transfer and referral to structural heart team, expedited pre-procedural CT, and a mini-multidisciplinary team comprising an interventional cardiologist, cardiac surgeon and imaging specialist. This streamlined approach reduced median admission-to-TAVR time from 22 to 10 days while numerically reducing a composite of death and acute kidney injury at 30 days (4.0% versus 20.7%; p=0.093) [17]. Wood et al.’s “Code TAVR” and Kitahara et al.’s “Door-to-Valve” offered additional practical recommendations including immediate structural heart team referral when severe AS with CS (SCAI stages C-E) is suspected, and establishing a pathway for prompt activation of hybrid catheterisation laboratories [7, 10].

Several factors may explain the inconsistent study findings. Two studies actively excluded haemodynamic instability on admission [17, 18], three were confined to cardiogenic shock cohorts [6, 19, 20], while the remainder enrolled broader ADAS populations. Presenting symptoms may reflect different structural pathologies. For example, syncope is associated with smaller LV cavity and reduced cardiac output, whereas dyspnoea may be due to advanced diastolic and/or systolic dysfunction [21]. Distinct haemodynamic patterns therefore suggest different initial stabilisation strategies, as cautious diuresis may alleviate congestion in dyspnoea but may further compromise cardiac output in syncope due to limited stroke-volume reserve.

Another reason for the null findings could be the failure to distinguish systems-related delays from clinically necessary delays. Clinical deterioration may prompt rescue intervention, increasing apparent risk in the early-intervention group. While delay may reflect modifiable constraints in referral, transfer, imaging and procedural capacity, it may also represent clinically necessary time to treat sepsis, neuroprognosticate, or stabilise another competing reversible illness. Finally, seven of eight studies excluded patients who deteriorated and died while awaiting TAVR or BAV, introducing selection and immortal time bias which favoured delayed intervention.

These findings have important implications for future research and service delivery. Prospective studies should enrol consecutive patients from hospital presentation, including those who deteriorate or die before intervention, and employ standardised definitions of ADAS, timing origin and clinical outcomes. Time-to-intervention analyses should account for time-dependent treatment allocation and competing risks, while distinguishing clinically indicated stabilisation from potentially avoidable systems delays. Plausible subgroups for prospective evaluation include patients with right ventricular dysfunction [22], syncope with preserved LVEF, and ADAS complicated by cardiogenic shock, incorporating serial SCAI staging to characterise haemodynamic trajectory. Ultimately, prospective pathway evaluations and, where feasible, randomised studies are needed to establish whether expedited intervention improves outcomes and whether clinically meaningful door-to-valve targets can be defined.

### Limitations

First, all included studies were observational with moderate to serious risk of bias, limiting causal inference. Second, heterogeneity in ADAS severity, intervention type, timing origin, early-treatment thresholds, recorded outcomes and follow-up limit comparability and clinical interpretation of pooled effects. Third, few studies contributed to each quantitative analysis, producing imprecise estimates and limiting assessment of publication bias.

## CONCLUSION

Current observational data do not yet establish an optimal door-to-valve intervention interval among patients with ADAS or a mortality benefit from expedited door-to-valve treatment.

Prospective and randomized-controlled trials on optimal valve intervention timing following ADAS are needed.

## ABBREVIATIONS

ADAS: Acute decompensated aortic stenosis
ASTRID-AS: Rapid Assessment and Treatment in Decompensated Aortic Stenosis
BAV: Balloon aortic valvuloplasty
CS: Cardiogenic shock
LVEF: Left ventricular ejection fraction
PCI: Percutaneous coronary intervention
SCAI: Society for Cardiovascular Angiography and Interventions
STEMI: ST-elevation myocardial infarction
TAVR: Transcatheter aortic valve replacement

## Data Availability Statement

All data underlying this article were extracted from previously published studies cited in the manuscript. The extracted study-level dataset and analytic code are available from the corresponding author upon reasonable request.

